# Casirivimab and imdevimab in patients admitted to hospital with COVID-19 (RECOVERY): a randomised, controlled, open-label, platform trial

**DOI:** 10.1101/2021.06.15.21258542

**Authors:** RECOVERY Collaborative Group, Peter W Horby, Marion Mafham, Leon Peto, Mark Campbell, Guilherme Pessoa-Amorim, Enti Spata, Natalie Staplin, Jonathan R Emberson, Benjamin Prudon, Paul Hine, Thomas Brown, Christopher A Green, Rahuldeb Sarkar, Purav Desai, Bryan Yates, Tom Bewick, Simon Tiberi, Tim Felton, J Kenneth Baillie, Maya H Buch, Lucy C Chappell, Jeremy N Day, Saul N Faust, Thomas Jaki, Katie Jeffery, Edmund Juszczak, Wei Shen Lim, Alan Montgomery, Andrew Mumford, Kathryn Rowan, Guy Thwaites, David M Weinreich, Richard Haynes, Martin J Landray

## Abstract

**Background:** REGEN-COV is a combination of 2 monoclonal antibodies (casirivimab and imdevimab) that bind to two different sites on the receptor binding domain of the SARS-CoV-2 spike protein. We aimed to evaluate the efficacy and safety of REGEN-COV in patients admitted to hospital with COVID-19.

**Methods:** In this randomised, controlled, open-label platform trial, several possible treatments were compared with usual care in patients hospitalised with COVID-19. Eligible and consenting patients were randomly allocated (1:1) to either usual standard of care alone (usual care group) or usual care plus a single dose of REGEN-COV 8g (casirivimab 4g and imdevimab 4g) by intravenous infusion (REGEN-COV group). The primary outcome was 28-day mortality assessed first among patients without detectable antibodies to SARS-CoV-2 at randomisation (seronegative) and then in the overall population. The trial is registered with ISRCTN (50189673) and clinicaltrials.gov (NCT04381936).

**Findings:** Between 18 September 2020 and 22 May 2021, 9785 patients were randomly allocated to receive usual care plus REGEN-COV or usual care alone, including 3153 (32%) seronegative patients, 5272 (54%) seropositive patients and 1360 (14%) patients with unknown baseline antibody status. In the primary efficacy population of seronegative patients, 396 (24%) of 1633 patients allocated to REGEN-COV and 451 (30%) of 1520 patients allocated to usual care died within 28 days (rate ratio 0·80; 95% CI 0·70-0·91; p=0·0010). In an analysis involving all randomised patients (regardless of baseline antibody status), 944 (20%) of 4839 patients allocated to REGEN-COV and 1026 (21%) of 4946 patients allocated to usual care died within 28 days (rate ratio 0·94; 95% CI 0·86-1·03; p=0·17). The proportional effect of REGEN-COV on mortality differed significantly between seropositive and seronegative patients (p value for heterogeneity = 0·001).

**Interpretation:** In patients hospitalised with COVID-19, the monoclonal antibody combination of casirivimab and imdevimab (REGEN-COV) reduced 28-day mortality among patients who were seronegative at baseline.

**Funding:** UK Research and Innovation (Medical Research Council) and National Institute of Health Research (Grant ref: MC_PC_19056).

## INTRODUCTION

Monoclonal antibodies (mAbs) are a set of identical antibodies that have high specificity and affinity for a single epitope. They have been demonstrated to be safe and effective in selected viral diseases when used for prophylaxis (respiratory syncytial virus) or treatment (Ebola virus disease).^1-3^ The clinical efficacy of mAbs in viral infections is thought to be mediated through direct binding to free virus particles and neutralisation of their ability to infect host cells. mAbs may also bind to viral antigens expressed on the surface of infected cells and stimulate antibody-dependent phagocytosis and cytotoxicity via the Fc portion of the mAb.^4^

SARS-CoV-2 infection is initiated by binding of the viral transmembrane spike glycoprotein to angiotensin converting enzyme 2 (ACE2) on the surface of host cells.^5^ The receptor binding domain of the spike glycoprotein is, consequently, the main target for neutralising antibodies.^6^ Following the emergence of SARS-COV-2, mAbs targeting the spike receptor binding domain were rapidly isolated from humanised mice and from peripheral B cells of recovered patients.^7,8^ Anti-SARS-CoV-2 spike protein neutralizing mAbs have demonstrated in vivo efficacy in both therapeutic and prophylactic settings in mouse, and non-human primates models, with decreases in viral load and lung pathology.^9-12^

Regeneron Pharmaceuticals (Tarrytown, New York, USA) has developed two non-competing, high-affinity human IgG1 anti-SARS-CoV-2 mAbs, casirivimab and imdevimab, which bind specifically to the receptor binding domain of the spike glycoprotein of SARS-CoV-2, blocking viral entry into host cells.^13^ A combination of antibodies that bind to non-overlapping epitopes, rather than a single antibody, is intended to minimize the likelihood of loss of antiviral activity due to naturally circulating viral variants or development of escape mutants under drug pressure.^14^ In a clinical study in non-hospitalised adults with SARS-COV-2 infection and risk factors for severe COVID-19, the combination of casirivimab and imdevimab (REGEN-COV) was safe and, compared to placebo, reduced virus load in the upper airway, shortened the time to symptom resolution, and reduced the composite outcome of COVID-19-related hospitalisation or all-cause mortality.^15,16^ Other anti-spike mAb products have also demonstrated an antiviral and clinical effect in non-hospitalised adults with SARS-COV-2 infection.^17,18^ In the United States, Emergency Use Authorization has been given for the use of bamlanivimab with etesevimab, REGEN-COV, and sotrovimab in non-hospitalised patients with mild to moderate COVID-19. The European Medicines Agency has authorised REGEN-COV for use in patients who are at high risk of progressing to severe COVID-19 but do not require supplemental oxygen. Interim results from a small trial of REGEN-COV in hospitalised patients requiring low-flow oxygen was consistent with a clinical benefit in seronegative patients.^19^

However, to date, no virus-directed therapy has been shown to reduce mortality in hospitalised patients with COVID-19, for whom the only treatments so far shown to reduce mortality have been those that modify the inflammatory response.^20-22^. The only published trial of an anti-spike mAb (bamlanivimab) in hospitalised patients was terminated for futility after 314 patients had been randomised.^23,24^ Two other studies of mAb products (VIR-7831 monotherapy, and BRII-196 with BRII-198 combination therapy) in hospitalized COVID-19 patients were also terminated for futility with sample sizes of 344 and 343 respectively.^25^ On first principles, the clinical response to antibody-based therapies may be greatest in individuals early in disease or who fail to mount an effective immune response. This is supported by evidence of clinical benefit in early disease and evidence that baseline anti-SARS-CoV-2 antibody status may be an important predictor of the effect of anti-spike mAbs on viral load.^15,16,19^ A significant proportion of hospitalised COVID-19 patients are seronegative on admission, and although a greater proportion already have detectable anti-SARS-CoV-2 antibodies, the quality of their immunological response may be poor since it has failed to prevent disease progression.^26^ As such, anti-spike mAbs may have benefit even in later COVID-19 disease. Here we report the results of a large randomised controlled trial of REGEN-COV in patients hospitalised with COVID-19.

## METHODS

### Study design and participants

The Randomised Evaluation of COVID-19 therapy (RECOVERY) trial is an investigator-initiated, individually randomised, controlled, open-label, platform trial to evaluate the effects of potential treatments in patients hospitalised with COVID-19. Details of the trial design and results for other possible treatments (dexamethasone, hydroxychloroquine, lopinavir-ritonavir, azithromycin, tocilizumab, convalescent plasma, colchicine and aspirin) have been published previously.^20,21,26-30^ The trial is underway at 177 hospitals in the United Kingdom supported by the National Institute for Health Research Clinical Research Network, two hospitals in Indonesia, and two hospitals in Nepal (appendix pp 3-27). Of these, 127 UK hospitals took part in the evaluation of REGEN-COV. The trial is coordinated by the Nuffield Department of Population Health at the University of Oxford (Oxford, UK), the trial sponsor. The trial is conducted in accordance with the principles of the International Conference on Harmonisation–Good Clinical Practice guidelines and approved by the UK Medicines and Healthcare products Regulatory Agency (MHRA) and the Cambridge East Research Ethics Committee (ref: 20/EE/0101). The protocol and statistical analysis plan are available in the appendix (pp 68-148) with additional information available on the study website www.recoverytrial.net.

Patients admitted to hospital were eligible for the study if they had clinically suspected or laboratory confirmed SARS-CoV-2 infection and no medical history that might, in the opinion of the attending clinician, put the patient at significant risk if they were to participate in the trial. Patients who had received intravenous immunoglobulin treatment during the current admission and children weighing <40 kg or aged <12 years were not eligible for randomisation to REGEN-COV. Pregnant or breastfeeding women were eligible for inclusion. Written informed consent was obtained from all patients, or a legal representative if patients were too unwell or unable to provide consent.

### Randomisation and masking

Baseline data were collected using a web-based case report form that included demographics, level of respiratory support, major comorbidities, suitability of the study treatment for a particular patient, and treatment availability at the study site (appendix pp 34-36).

Baseline presence of anti-SARS-CoV-2 antibodies was to be determined for each participant using serum samples taken at the time of randomisation. Analysis was done at a central laboratory with a validated 384 well plate indirect ELISA for anti-spike IgG (appendix p 28).^31^ Participants were categorised as seropositive or seronegative using a predefined assay threshold with a 99% or higher sensitivity and specificity in detecting individuals with SARS-CoV-2 infection at least 20 days previously.^31^

Eligible and consenting patients were assigned in a 1:1:1 ratio to either usual standard of care, usual standard of care plus REGEN-COV or usual standard of care plus convalescent plasma (until 15 January 2021), using web-based simple (unstratified) randomisation with allocation concealed until after randomisation (appendix pp 32-33). For some patients, REGEN-COV was unavailable at the hospital at the time of enrolment or was considered by the managing physician to be either definitely indicated or definitely contraindicated. These patients were excluded from the randomised comparison between REGEN-COV and usual care. Patients allocated to REGEN-COV were to receive a single dose of REGEN-COV 8g (casirivimab 4g and imdevimab 4g) in 250ml 0.9% saline infused intravenously over 60 minutes +/- 15 minutes as soon as possible after randomisation.

As a platform trial, and in a factorial design, patients could be simultaneously randomised to other treatment groups: i) azithromycin versus usual care, ii) colchicine versus usual care, iii) aspirin versus usual care, and iv) baricitinib versus usual care. Further details of when these factorial randomisations were open is provided in the supplementary appendix (pp 32-33). Until 24 January 2021, the trial also allowed a subsequent randomisation for patients with progressive COVID-19 (evidence of hypoxia and a hyper-inflammatory state) to tocilizumab versus usual care. Participants and local study staff were not masked to the allocated treatment. The trial steering committee, investigators, and all other individuals involved in the trial were masked to outcome data during the trial.

### Procedures

Early safety outcomes were recorded by site staff using an online form 72 hours after randomisation (appendix pp 37–41). An online follow-up form was completed by site staff when patients were discharged, had died, or at 28 days after randomisation, whichever occurred first (appendix pp 42–48). Information was recorded on adherence to allocated trial treatment, receipt of other COVID-19 treatments, duration of admission, receipt of respiratory or renal support, and vital status (including cause of death). In addition, routine health-care and registry data were obtained, including information on vital status at day 28 (with date and cause of death); discharge from hospital; and receipt of respiratory support or renal replacement therapy.

### Outcomes

Outcomes were assessed at 28 days after randomisation, with further analyses specified at 6 months. The primary outcome was 28-day all-cause mortality. Secondary outcomes were time to discharge from hospital, and, among patients not on invasive mechanical ventilation at randomisation, the composite outcome of invasive mechanical ventilation (including extra-corporeal membrane oxygenation) or death. Prespecified subsidiary clinical outcomes were use of invasive or non-invasive ventilation among patients not on any ventilation at randomisation, time to successful cessation of invasive mechanical ventilation (defined as cessation of invasive mechanical ventilation within, and survival to, 28 days), and use of renal dialysis or haemofiltration. Information on suspected serious adverse reactions was collected in an expedited fashion to comply with regulatory requirements. Details of the methods used to ascertain and derive outcomes are provided in the appendix (pp.149-169).

Prespecified safety outcomes were cause-specific mortality, major cardiac arrhythmia, and thrombotic and major bleeding events (only collected since 6 November 2021). Information on early safety outcomes at 72 h following randomisation (worsening respiratory status, severe allergic reactions, fever, sudden hypotension, clinical haemolysis, and thrombotic events) ceased on 19 February 2021 on the advice of the Data Monitoring Committee and in accordance with the protocol.

### Statistical Analysis

For all outcomes, intention-to-treat analyses compared patients randomised to REGEN-COV with patients randomised to usual care but for whom REGEN-COV was both available and suitable as a treatment. For the primary outcome of 28-day mortality, the log-rank observed minus expected statistic and its variance were used to both test the null hypothesis of equal survival curves (i.e., the log-rank test) and to calculate the one-step estimate of the average mortality rate ratio. We constructed Kaplan-Meier survival curves to display cumulative mortality over the 28-day period.

For this preliminary report, information on the primary outcome is available for 99% of randomised patients. Those patients who had not been followed for 28 days and were not known to have died by the time of the data cut for this preliminary analysis (25 May 2021) were either censored on 25 May 2021 or, if they had already been discharged alive, were right-censored for mortality at day 29 (that is, in the absence of any information to the contrary they were assumed to have survived 28 days). [Note: This censoring rule will not be necessary when all patients have completed the 28 day follow-up period on 19 June 2021.]

We used the same method to analyse time to hospital discharge and successful cessation of invasive mechanical ventilation, with patients who died in hospital right-censored on day 29. Median time to discharge was derived from Kaplan-Meier estimates. For the pre-specified composite secondary outcome of progression to invasive mechanical ventilation or death within 28 days (among those not receiving invasive mechanical ventilation at randomisation), and the subsidiary clinical outcomes of receipt of ventilation and use of haemodialysis or haemofiltration, the precise dates were not available and so the risk ratio was estimated instead. Estimates of rate and risk ratios (both denoted RR) are shown with 95% confidence intervals.

In the light of new evidence which became available during the trial, it was hypothesised that any beneficial effect of REGEN-COV would be larger among seronegative participants (and may be negligible in seropositive participants).^15,19^ Consequently, prior to any unblinding of results, the trial steering committee specified that hypothesis-testing of the effect of allocation to REGEN-COV on 28-day mortality (and secondary outcomes) would first be done only in seronegative participants (appendix pp. 142-144). Hypothesis testing of the primary outcome among all randomised patients was then only to be done if a reduction in mortality in seronegative patients was seen at 2P<0.05. A prespecified comparison of the effects of allocation to REGEN-COV on 28-day mortality in seronegative versus seropositive participants was done by performing a test for heterogeneity. Tests for heterogeneity according to other baseline characteristics (age, sex, ethnicity, level of respiratory support, days since symptom onset, and use of corticosteroids) (appendix p 133-134) were also prespecified.

The full database is held by the study team which collected the data from study sites and performed the analyses at the Nuffield Department of Population Health, University of Oxford (Oxford, UK).

As stated in the protocol, appropriate sample sizes could not be estimated when the trial was being planned at the start of the COVID-19 pandemic. On 27 April 2021, the trial steering committee, whose members were unaware of the results of the trial comparisons, determined that, with over 9700 patients recruited to the REGEN-COV comparison and average daily recruitment of 4 patients, further recruitment was unlikely to increase reliability of the results materially so should discontinue (appendix p 33-34). The statistical analysis plan was finalised and published on 21 May 2021 (without any knowledge of the study results) (appendix pp 112-148) and recruitment to the REGEN-COV comparison was closed on 22 May 2021. The trial steering committee and all other individuals involved in the trial were masked to outcome data until after the close of recruitment (appendix p 49).

Analyses were performed using SAS version 9.4 and R version 4.0.3. The trial is registered with ISRCTN (50189673) and clinicaltrials.gov (NCT04381936).

## RESULTS

Between 18 September 2020 and 22 May 2021, 11464 (47%) of 24343 patients enrolled into the RECOVERY trial at one of the 127 sites were eligible to be randomly allocated to REGEN-COV (i.e. REGEN-COV was available in the hospital at the time and the attending clinician was of the opinion that the patient had no known indication for or contraindication to REGEN-COV, figure 1). 4839 patients were randomly allocated to REGEN-COV and 4946 were randomly allocated to usual care. The mean age of study participants in this comparison was 61.9 years (SD 14.5) and the median time since symptom onset was 9 days (IQR 6 to 12 days) (webtable 1). At randomisation, 9169 (94%) patients were receiving corticosteroids. 5272 (54%) were seropositive at baseline, 3153 (32%) were seronegative, and serostatus was unknown for 1360 (14%) (table 1, webtables 1 and 2).

**Table 1:** Baseline characteristics (seronegative and all participants) by treatment allocation.

|  | Seronegative patients |  | All patients |  |
| --- | --- | --- | --- | --- |
|  | REGEN-COV<br>(n=1633) | Usual Care<br>(n=1520) | REGEN-COV<br>(n=4839) | Usual Care<br>(n=4946) |
| Age, years | 63.2 (15.5) | 64.0 (15.2) | 61.9 (14.6) | 61.9 (14.4) |
| <70* | 1054 (65) | 943 (62) | 3389 (70) | 3454 (70) |
| 70 to 79 | 348 (21) | 344 (23) | 936 (19) | 962 (19) |
| ≥80 | 231 (14) | 233 (15) | 514 (11) | 530 (11) |
| Sex |  |  |  |  |
| Men | 995 (61) | 879 (58) | 3033 (63) | 3095 (63) |
| Women† | 638 (39) | 641 (42) | 1806 (37) | 1851 (37) |
| Ethnicity |  |  |  |  |
| White | 1324 (81) | 1250 (82) | 3768 (78) | 3810 (77) |
| Black, Asian, and minority ethnic | 147 (9) | 136 (9) | 588 (12) | 696 (14) |
| Unknown | 162 (10) | 134 (9) | 483 (10) | 440 (9) |
| Number of days since symptom onset | 7 (4-10) | 7 (5-9) | 9 (6-12) | 9 (6-12) |
| Number of days since admission to hospital | 1 (1-2) | 1 (1-3) | 2 (1-3) | 2 (1-3) |
| Respiratory support received |  |  |  |  |
| No oxygen received | 182 (11) | 148 (10) | 332 (7) | 309 (6) |
| Simple oxygen | 1085 (66) | 995 (65) | 2980 (62) | 3016 (61) |
| Non-invasive ventilation | 332 (20) | 341 (22) | 1244 (26) | 1317 (27) |
| Invasive mechanical ventilation | 34 (2) | 36 (2) | 283 (6) | 304 (6) |
| Previous diseases |  |  |  |  |
| Diabetes | 403 (25) | 407 (27) | 1240 (26) | 1337 (27) |
| Heart disease | 407 (25) | 398 (26) | 1038 (21) | 1061 (21) |
| Chronic lung disease | 455 (28) | 458 (30) | 1085 (22) | 1159 (23) |
| Tuberculosis | 7 (<1) | 5 (<1) | 18 (<1) | 16 (<1) |
| HIV | 7 (<1) | 4 (<1) | 24 (<1) | 22 (<1) |
| Severe liver disease‡ | 28 (2) | 17 (1) | 69 (1) | 70 (1) |
| Severe kidney impairment§ | 114 (7) | 114 (8) | 266 (5) | 242 (5) |
| Any of the above | 935 (57) | 913 (60) | 2557 (53) | 2662 (54) |
| SARS-CoV-2 PCR test result |  |  |  |  |
| Positive | 1580 (97) | 1470 (97) | 4680 (97) | 4791 (97) |
| Negative | 17 (1) | 16 (1) | 38 (1) | 53 (1) |
| Unknown | 36 (2) | 34 (2) | 121 (3) | 102 (2) |
| Patient SARS-CoV-2 antibody test result |  |  |  |  |
| Positive | 0 | 0 | 2636 (54) | 2636 (53) |
| Negative | 1633 (100) | 1520 (100) | 1633 (34) | 1520 (31) |
| Missing | 0 | 0 | 570 (12) | 790 (16) |
| Corticosteroids received |  |  |  |  |
| Yes | 1481 (91) | 1399 (92) | 4530 (94) | 4639 (94) |
| No | 152 (9) | 118 (8) | 308 (6) | 299 (6) |
| Not recorded | 0 | 3 (<1) | 1 (<1) | 8 (<1) |
| Other randomised treatments |  |  |  |  |
| Azithromycin | 38 (2) | 43 (3) | 124 (3) | 124 (3) |
| Colchicine | 364 (22) | 350 (23) | 1085 (22) | 1139 (23) |
| Aspirin | 405 (25) | 372 (24) | 1339 (28) | 1389 (28) |

Data are mean (SD), n (%), or median (IQR). \*Includes 11 children (<18 years). † Includes 25 pregnant women. ‡ Defined as requiring ongoing specialist care. § Defined as estimated glomerular filtration rate <30 mL/min per 1.73 m<sup>2</sup>

**Table 2:** Effect of allocation to REGEN-COV on key study outcomes among seronegative participants.

|  | REGEN-COV<br>(n=1633) | Usual Care<br>(n=1520) | RR (95% CI) |
| --- | --- | --- | --- |
| **Primary outcome** |  |  |  |
| Mortality at 28 days | 396 (24%) | 451 (30%) | 0.80 (0.70-0.91) |
| **Secondary outcomes** |  |  |  |
| Median duration of hospitalisation, days | 13 (7 to >28) | 17 (7 to >28) | - |
| Discharged from hospital within 28 days | 1046 (64%) | 878 (58%) | 1.19 (1.08-1.30) |
| Invasive mechanical ventilation or death* | 487/1599 (30%) | 542/1484 (37%) | 0.83 (0.75-0.92) |
| Invasive mechanical ventilation | 189/1599 (12%) | 200/1484 (13%) | 0.88 (0.73-1.06) |
| Death | 383/1599 (24%) | 434/1484 (29%) | 0.82 (0.73-0.92) |
| **Subsidiary outcomes** |  |  |  |
| Use of ventilation † | 355/1267 (28%) | 370/1143 (32%) | 0.87 (0.77-0.98) |
| Non-invasive ventilation | 341/1267 (27%) | 360/1143 (31%) | 0.85 (0.75-0.97) |
| Invasive mechanical ventilation | 89/1267 (7%) | 119/1143 (10%) | 0.67 (0.52-0.88) |
| Successful cessation of invasive mechanical ventilation ‡ | 9/34 (26%) | 12/36 (33%) | 0.86 (0.36-2.03) |
| Renal replacement therapy § | 68/1616 (4%) | 64/1498 (4%) | 0.98 (0.71-1.38) |

Data are n (%). median (IQR) or n/N (%). RR=rate ratio for the outcomes of 28-day mortality, hospital discharge, and successful cessation of invasive mechanical ventilation, and risk ratio for other outcomes.
\* Analyses exclude those on invasive mechanical ventilation at randomisation.
† Analyses exclude those on invasive or non-invasive ventilation at randomisation.
‡ Analyses exclude those not receiving invasive mechanical ventilation at randomisation.
§ Analyses exclude those on renal replacement therapy at randomisation.

**Figure 1:**
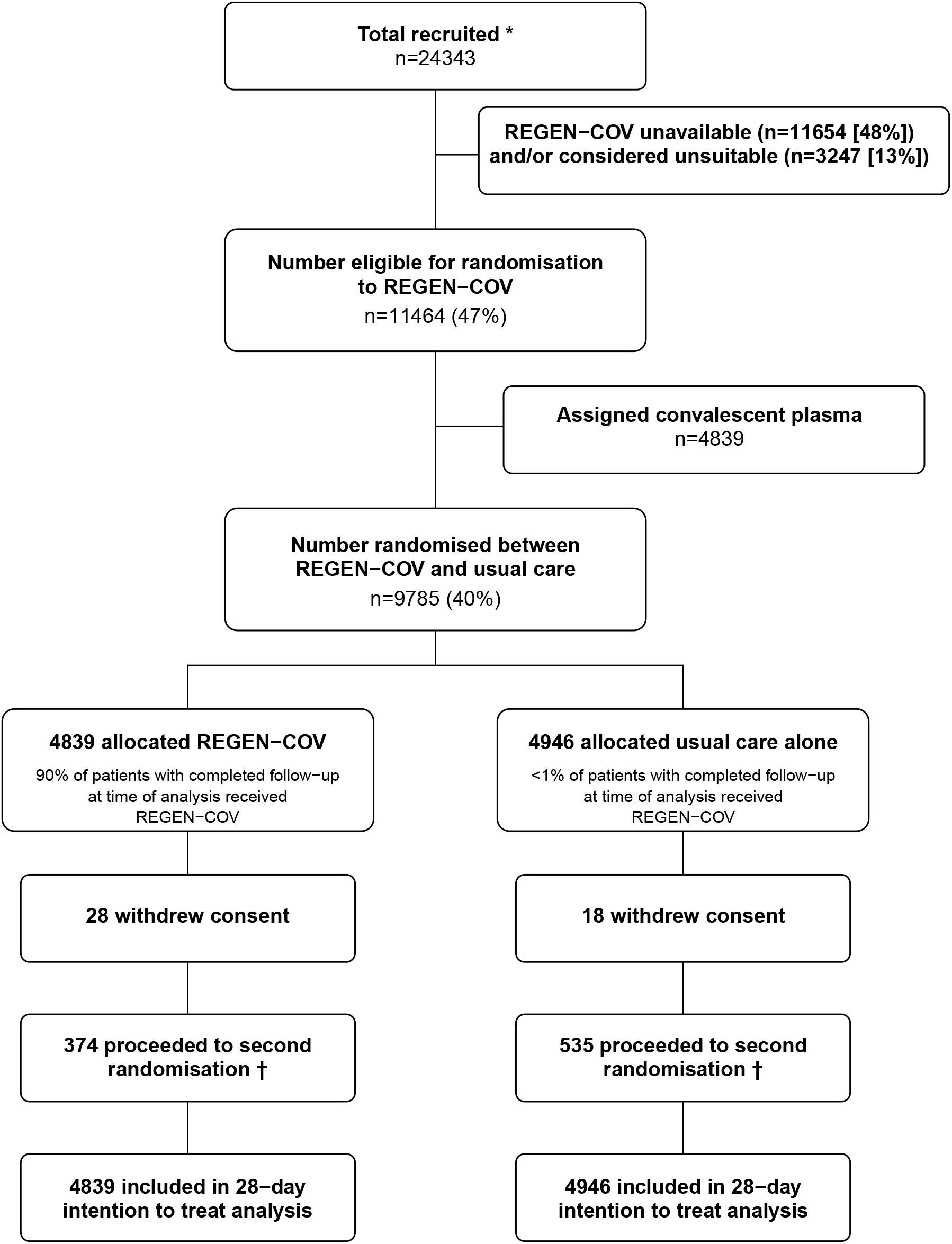
Trial profile. ITT=intention to treat. * Number recruited overall during period that adult participants could be recruited into REGEN-COV comparison. Of the 9785 randomised to REGEN-COV vs usual care, 4535 were additionally randomised to colchicine vs usual care (2238 [46%] of the REGEN-COV group vs 2297 [46%] of the usual care group); 5507 were additionally randomised to aspirin vs usual care (2665 [55%] of the REGEN-COV group vs 2842 [57%] of the usual care group), and 1772 patients were additionally randomised to baricitinib vs usual care (889 [18%] of the REGEN-COV group vs 883 [18%] of the usual care group). † Includes 185/4839 (4%) patients in the REGEN-COV arm and 271/4946 (5%) patients in the usual care arm allocated to tocilizumab.

The follow-up form was completed for 4773 (99%) in the REGEN-COV group and 4899 (99%) in the usual care group. Among patients with a completed follow-up form, 90% allocated to REGEN-COV received the treatment compared with <1% allocated to usual care (figure 1). Use of other treatments for COVID-19 was similar among patients allocated REGEN-COV and among those allocated usual care, with about one-quarter receiving remdesivir and one-seventh receiving tocilizumab (webtable 3). Primary and secondary outcome data are known for 99% of randomly assigned patients.

Among patients who were known to be seronegative at baseline, allocation to REGEN-COV was associated with a significant reduction in the primary outcome of 28-day mortality compared with usual care alone: 396 (24%) of 1633 patients in the REGEN-COV group died vs 451 (30%) of 1520 patients in the usual care group (rate ratio 0·80; 95% CI, 0·70–0·91; p=0·0010; table 2, figure 2a, and figure 3). The proportional effect of REGEN-COV on mortality differed significantly between seropositive and seronegative patients (test for heterogeneity p=0.001; figure 3). Among all patients randomised (including those with negative, positive, or unknown baseline antibody status), there was no significant difference in the primary outcome of 28-day mortality between the two randomised groups: 944 (20%) of 4839 patients in the REGEN-COV group died vs. 1026 (21%) of 4946 patients in the usual care group (rate ratio 0·94; 95% CI, 0·86 to 1·03; p=0·17; webtable 4, figure 2b, and figure 3).

**Figure 2:**
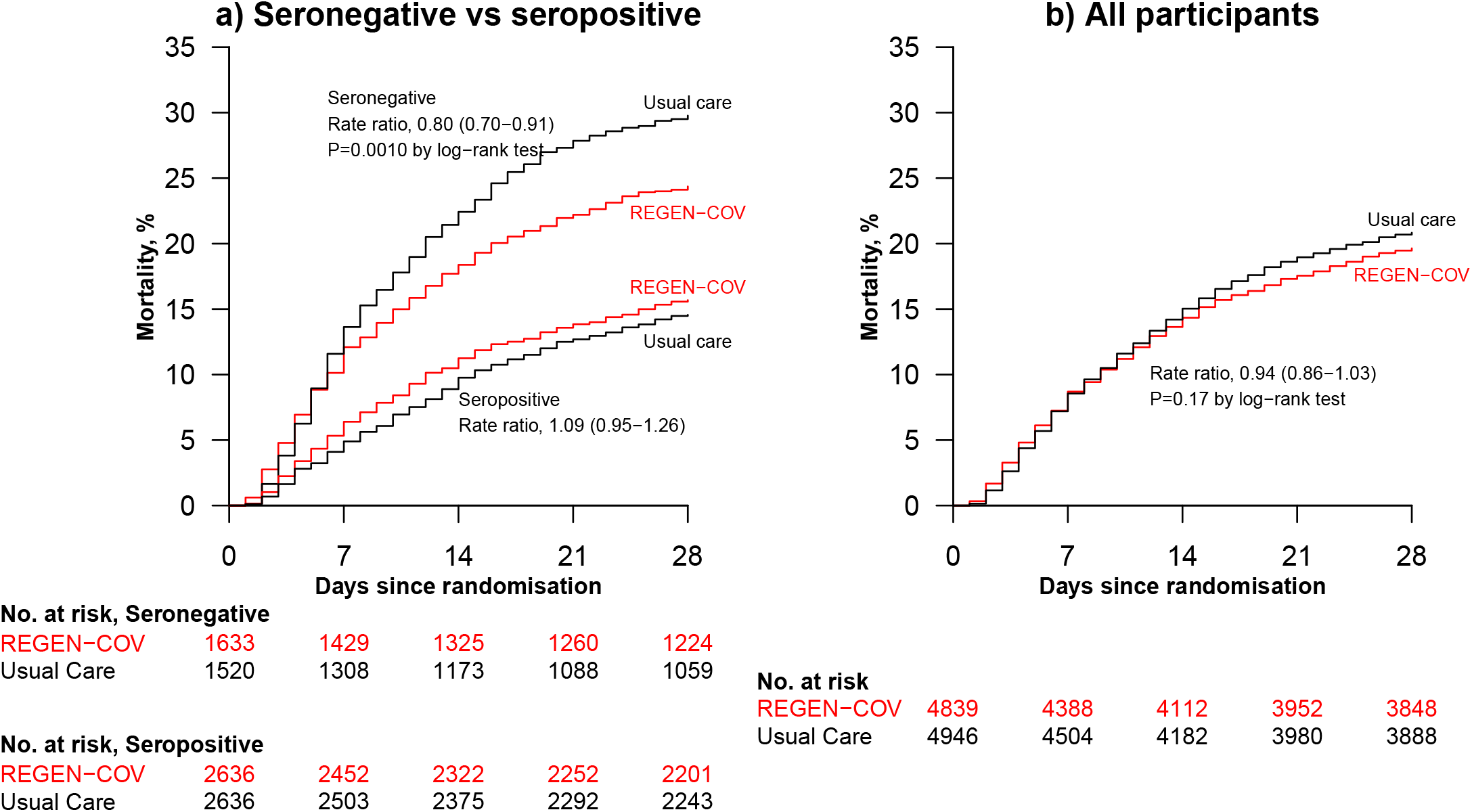
Effect of allocation to REGEN-COV on 28-day mortality (a) in seronegative patients and seropositive patients (b) overall.

**Figure 3:**
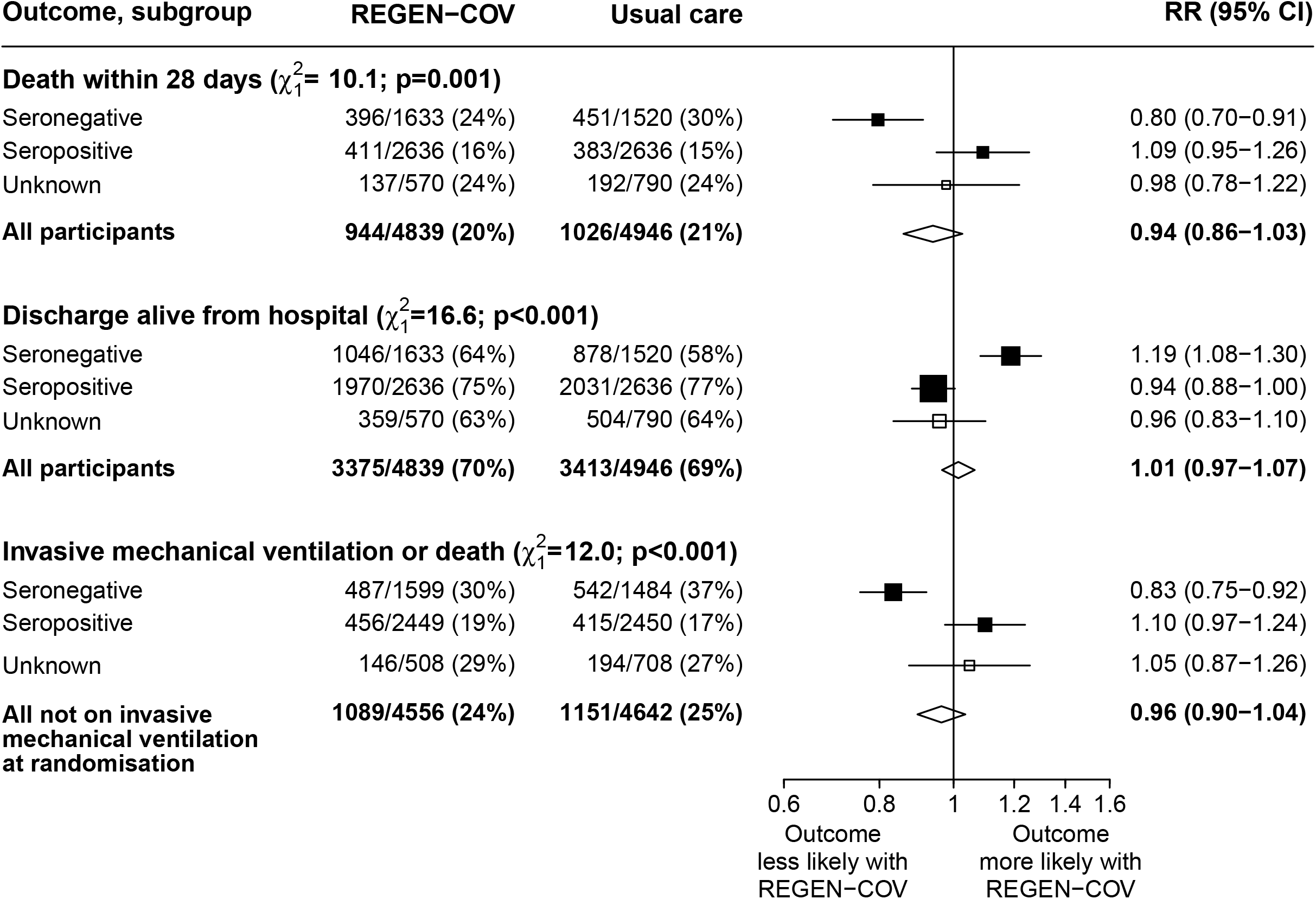
Primary and secondary outcomes, overall and by baseline antibody status. Subgroup–specific rate ratio estimates are represented by squares (with areas of the squares proportional to the amount of statistical information) and the lines through them correspond to the 95% CIs. The tests for heterogeneity compare the log RRs in the seronegative versus seropositive subgroups (ie, ignoring those with unknown antibody status).

In both the seronegative patients and in all patients combined, the proportional effects on mortality seen in the respective populations were consistent across all other pre-specified subgroups (webfigure 1 and webfigure 2). Results were virtually identical when restricted to participants with a positive SARS-CoV-2 PCR test (webtable 5). In a sensitivity analysis using a Cox model adjusted for all pre-specified subgroups, allocation to REGEN-COV was associated with a mortality rate ratio of 0·85 (95% CI 0·74-0·98) in seronegative patients (webtable 5). Among all participants, there was no evidence that the effect on mortality varied depending on concurrent randomised allocation to azithromycin, colchicine, or aspirin (all interaction p-values >0.1).

Among seronegative patients, discharge alive within 28 days was more common among those allocated to REGEN-COV compared with usual care (64% vs. 58%; rate ratio 1·19, 95% CI 1·08 to 1·30; median 13 days [IQR 7 to >28] vs. 17 days [IQR 7 to >28]) (table 2, figure 3 and webfigure 3a). However, there was no meaningful difference among the overall study population (70% vs. 69%; rate ratio 1·01, 95% CI 0·97 to 1·07; median 10 days [IQR 6 to >28] vs. 10 days [IQR 5 to >28]) (webtable 4, figure 3 and webfigure 3b).

Among seronegative patients not on invasive mechanical ventilation at baseline, allocation to REGEN-COV was associated with a lower risk of progressing to the composite secondary outcome of invasive mechanical ventilation or death (30% vs. 37%, risk ratio 0·83, 95% CI 0·75 to 0·92) (table 2 and figure 3). However, there was no difference among the overall study population (24% vs. 25%, risk ratio 0·96, 95% CI 0·90 to 1·04) (webtable 4 and figure 3).

There was clear evidence that the proportional effects on each of these secondary outcomes differed significantly between seropositive and seronegative patients (p value for heterogeneity both <0.001) (figure 3). There was no good evidence of differences in treatment effect in other subgroups of patients (webfigures 4 and 5).

Among seronegative patients, allocation to REGEN-COV versus usual care was associated with less frequent progression to use of ventilation among patients not on such treatment at baseline versus usual care (28% vs 32%; risk ratio 0·87, 95% CI 0·77 to 0·98) (table 2) but not in the overall study population (23% vs. 24%; risk ratio 0·95, 95% CI 0·87 to 1·04) (webtable 4). There were no meaningful differences in progression to renal replacement therapy, non-COVID mortality, cardiac arrhythmia, thrombosis or major bleeding either in the seronegative or overall study populations (table 2, webtables 4, 6, 7 and 8).

Information on potential infusion reactions occurring within the first 72 hours after randomisation was collected for 1792 patients in the REGEN-COV group and 1714 patients in the usual care group (before collection of these data stopped on 19 February 2021): The reported frequency of fever (4% vs. 3%), sudden hypotension (4% vs. 2%), and thrombotic events (2% vs. 1%) was marginally higher in the REGEN-COV group vs. the usual care group while the frequency of sudden worsening in respiratory status (21% vs. 22%) and clinical haemolysis (1% vs. 2%) was marginally lower (webtable 9). There were 5 reports of a serious adverse reaction believed to be related to REGEN-COV (webtable 10).

## DISCUSSION

In this large, randomised trial, allocation to REGEN-COV in patients who were anti-SARS-CoV-2 antibody negative at randomisation significantly reduced 28-day mortality by about one-fifth, an absolute benefit of 6 fewer deaths per 100 patients allocated REGEN-COV. In addition, allocation to REGEN-COV was associated with an increased rate of discharge alive from hospital within the first 28 days and a reduced rate of progression to invasive mechanical ventilation or death in these patients. By contrast, no such benefits were seen for patients who were anti-SARS-CoV-2 antibody positive at randomisation. Consequently, when all patients were considered together (including those with unknown antibody status), allocation to REGEN-COV was associated with non-significant differences in clinical outcomes. Only one other trial has reported the effects of an anti-spike mAb in hospitalised COVID-19 patients, and this trial was terminated for futility based on clinical status at day 5 in 314 patients.^23^ However, the result were not reported by baseline serostatus and that trial was underpowered to detect moderate effects in sub-groups. Whilst two other trials of mAbs in hospitalised patients were also terminated for futility by the same group, full details are not yet published.^25^

Based on our findings, any therapeutic use of REGEN-COV in the hospital setting may be best restricted to seronegative patients. This would require serological testing prior to drug administration. High-performance, laboratory-based commercial assays for SARS-CoV-2 antibodies are available and used in high-income healthcare settings. However, they are not widely available in lower income settings.^32^ Point-of-care lateral-flow immunoassays have been developed but some have suboptimal performance and their suitability for guiding therapeutic decisions, as opposed to sero-epidemiological studies, requires further evaluation.^31,33,34^ Assays with lower costs and technological requirements than commercial bench-top systems and better performance than lateral-flow immunoassays have been developed and may offer more scalable and affordable options for serostatus evaluation but these also require further evaluation before clinical use.^35^

In October 2020 the independent data monitoring committee of an industry sponsored trial of REGEN-COV in hospitalised COVID-19 patients recommended that recruitment of patients on high-flow oxygen or mechanical ventilation be suspended because of a potential safety signal.^36^ However, we did not observe any evidence that the proportional effect of REGEN-COV on mortality varied by level of respiratory support received at randomisation, either when assessed in all participants or when assessed only in the sub-group of seronegative participants.

mAbs are susceptible to the evolution of viral resistance if substitutions in the targeted epitope reduce or abrogate antibody binding, and an Emergency Use Authorisation for monotherapy with the mAb LY-CoV555 was revoked due to resistance in several major virus variants.^37^ This risk can be reduced by using a combination of mAbs that bind to non-overlapping epitopes.^14^ Whilst we did not study the emergence of resistance variants in this trial, the major variants circulating in the UK throughout the trial, including B.1.1.7 (alpha) variant which was the dominant variant in the UK from December 2020 to April 2021, remained sensitive to REGEN-COV.^38,39^ Although spike glycoprotein mutations in some variants (e.g. B.1.351 [beta] and B.1.617 [delta]) have been associated with a reduction of neutralisation activity of casivirimab, the combination of casirivimab with imdevimab retains potency against these variants due to the inhibitory activity of imdevimab.^38-41^ However, continued monitoring of resistance patterns is imperative to detect variants with resistance to both components.

Strengths of this trial included that it was randomised, had a large sample size, broad eligibility criteria and more than 99% of patients were followed up for the primary outcome. Information on virological outcomes was not collected, nor was information on radiological or physiological outcomes. Although this randomised trial is open label (i.e., participants and local hospital staff are aware of the assigned treatment), the outcomes are unambiguous and were ascertained without bias through linkage to routine health records. The dose of REGEN-COV used in this study was high compared to those used in outpatient studies; understanding the effects of lower doses would require additional evidence from a randomized controlled trial.^16^

In summary, this large, randomised trial provides the first evidence that an antiviral therapy can reduce mortality in hospitalised COVID-19 patients and the results support the use of REGEN-COV in seronegative patients hospitalised with COVID-19.

## Supporting information

CONSORT Checklist

## Data Availability

The protocol, consent form, statistical analysis plan, definition & derivation of clinical characteristics & outcomes, training materials, regulatory documents, and other relevant study materials are available online at www.recoverytrial.net. As described in the protocol, the trial Steering Committee will facilitate the use of the study data and approval will not be unreasonably withheld. Deidentified participant data will be made available to bona fide researchers registered with an appropriate institution within 3 months of publication. However, the Steering Committee will need to be satisfied that any proposed publication is of high quality, honours the commitments made to the study participants in the consent documentation and ethical approvals, and is compliant with relevant legal and regulatory requirements (e.g. relating to data protection and privacy). The Steering Committee will have the right to review and comment on any draft manuscripts prior to publication. Data will be made available in line with the policy and procedures described at: https://www.ndph.ox.ac.uk/data-access. Those wishing to request access should complete the form at
https://www.ndph.ox.ac.uk/files/about/data_access_enquiry_form_13_6_2019.docx
and e-mailed to

https://www.ndph.ox.ac.uk/data-access

## Data Availability

https://www.ndph.ox.ac.uk/data-access

## Contributors

This manuscript was initially drafted by the PWH and MJL, further developed by the Writing Committee, and approved by all members of the trial steering committee. PWH and MJL vouch for the data and analyses, and for the fidelity of this report to the study protocol and data analysis plan. PWH, MM JKB, MB, LCC, JD, SNF, TJ, EJ, KJ, WSL, AMo, AMu, KR, RH, and MJL designed the trial and study protocol. MM, LP, MC, G P-A, BP, PH, TB, CAG, RS, PD, BY, TB, ST, TF, and the Data Linkage team at the RECOVERY Coordinating Centre, and the Health Records and Local Clinical Centre staff listed in the appendix collected the data. ES, NS, and JRE did the statistical analysis. All authors contributed to data interpretation and critical review and revision of the manuscript. PWH and MJL had access to the study data and had final responsibility for the decision to submit for publication.

## Writing Committee (on behalf of the RECOVERY Collaborative Group)

Peter W Horby,^*^ Marion Mafham,^*^ Leon Peto, Mark Campbell, Guilherme Pessoa-406 Amorim, Enti Spata, Natalie Staplin, Jonathan R Emberson, Benjamin Prudon, Paul Hine, Thomas Brown, Christopher A Green, Rahuldeb Sarkar, Purav Desai, Bryan Yates, Tom Bewick, Simon Tiberi, Tim Felton, J Kenneth Baillie, Maya H Buch, Lucy C Chappell, Jeremy Day, Saul N Faust, Thomas Jaki, Katie Jeffery, Edmund Juszczak, Wei Shen Lim, Alan Montgomery, Andrew Mumford, Kathryn Rowan, Guy Thwaites, David M Weinreich, Richard Haynes,^+^ Martin J Landray.^+^

^*^PWH and MM made an equal contribution

^+^RH and MJL made an equal contribution

## Data Monitoring Committee

Peter Sandercock, Janet Darbyshire, David DeMets, Robert Fowler, David Lalloo, Mohammed Munavvar (from January 2021), Ian Roberts (until December 2020), Adilia Warris (from March 2021), Janet Wittes.

## Data sharing

The protocol, consent form, statistical analysis plan, definition & derivation of clinical characteristics & outcomes, training materials, regulatory documents, and other relevant study materials are available online at www.recoverytrial.net. As described in the protocol, the trial Steering Committee will facilitate the use of the study data and approval will not be unreasonably withheld. Deidentified participant data will be made available to bona fide researchers registered with an appropriate institution within 3 months of publication. However, the Steering Committee will need to be satisfied that any proposed publication is of high quality, honours the commitments made to the study participants in the consent documentation and ethical approvals, and is compliant with relevant legal and regulatory requirements (e.g. relating to data protection and privacy). The Steering Committee will have the right to review and comment on any draft manuscripts prior to publication. Data will be made available in line with the policy and procedures described at: https://www.ndph.ox.ac.uk/data-access. Those wishing to request access should complete the form at https://www.ndph.ox.ac.uk/files/about/data_access_enquiry_form_13_6_2019.docx and e-mailed to

## Acknowledgements

Above all, we would like to thank the thousands of patients who participated in this trial. We would also like to thank the many doctors, nurses, pharmacists, other allied health professionals, and research administrators at 177 NHS hospital organisations across the whole of the UK, supported by staff at the National Institute of Health Research (NIHR) Clinical Research Network, NHS DigiTrials, Public Health England, Department of Health & Social Care, the Intensive Care National Audit & Research Centre, Public Health Scotland, National Records Service of Scotland, the Secure Anonymised Information Linkage (SAIL) at University of Swansea, NHS Blood & Transplant and the NHS in England, Scotland, Wales and Northern Ireland.

The RECOVERY trial is supported by grants to the University of Oxford from UK Research and Innovation (UKRI) and NIHR (MC_PC_19056), the Wellcome Trust (Grant Ref: 222406/Z/20/Z) through the COVID-19 Therapeutics Accelerator, and by core funding provided by the NIHR Oxford Biomedical Research Centre, the Wellcome Trust, the Bill and Melinda Gates Foundation, the Foreign, Commonwealth and Development Office, Health Data Research UK, the Medical Research Council Population Health Research Unit, the NIHR Health Protection Unit in Emerging and Zoonotic Infections, and NIHR Clinical Trials Unit Support Funding. TJ is supported by a grant from UK Medical Research Council (MC_UU_00002/14) and an NIHR Senior Research Fellowship (NIHR-SRF-2015-08-001). WSL is supported by core funding provided by NIHR Nottingham Biomedical Research Centre. Tocilizumab was provided free of charge for this trial by Roche Products Limited. Regeneron Pharmaceuticals supported the trial through provision of REGEN-COV. The views expressed in this publication are those of the authors and not necessarily those of the NHS, the NIHR, the Department of Health and Social Care, or Regeneron Pharmaceuticals.

